# Effect of social media mental health messaging on mental help-seeking behaviors in the sub-Saharan African population: a systematic review protocol

**DOI:** 10.1101/2025.10.02.25337221

**Authors:** Priscilla Aboagyewaa Boateng, Isaiah Osei Duah, Josephine Ampong, Margaret Dowuona-Hammond, Laura Marciano

## Abstract

Mental health is a major public health concern, with disproportionate burdens in sub-Saharan Africa (SSA), where access to care is limited and stigma remains high. Social media platforms such as Facebook, WhatsApp, X, YouTube, TikTok, and other digital platforms provide opportunities for rapid information sharing, peer engagement, and tailored interventions that may enhance literacy and normalize help-seeking. Yet, they also pose risks, including misinformation, exposure to harmful content, and reinforcement of stigma within culturally diverse, community-based contexts. Despite this potential, evidence from SSA on the effects of social media messaging on mental health knowledge, attitudes, behaviors, and help-seeking is scarce, as most studies focus on prevalence or barriers to care. This systematic review will assess the effect of social media mental health messaging on mental-help-seeking behaviors in SSA. Electronic databases, including PubMed/MEDLINE, Psychological Information Database (PsycINFO), Cumulative Index to Nursing and Allied Health Literature (CINAHL), Communication and Mass Media Complete, Scopus, Cochrane Central Register of Controlled Trials (CENTRAL), Web of Science Core Collection, Educational Resource Information Centre (ERIC), ProQuest Sociology, and Medline via ProQuest will be systematically queried using predefined keywords. Eligible studies will include those examining the use of online mental health-related messages in SSA, analyzing audience engagement, behavioral and psychological outcomes, and any kind of intervention studies. Methodological quality and risk of bias will be assessed using validated tools appropriate to each study design, including the Cochrane Risk of Bias 2 (RoB 2) tool for randomized trials, the Risk of Bias in Non-Randomized Studies of Interventions (ROBINS-I), and the Strengthening the Reporting of Observational Studies in Epidemiology (STROBE) checklist for observational studies. Where appropriate, data will be synthesized with or without meta-analysis. Collectively, this synthesis will clarify how social media shapes mental health outcomes, identify gaps, and inform culturally sensitive interventions to improve communication, reduce disparities, and promote mental health help-seeking in SSA.

## Introduction

Mental health is a major public health concern, with a notable resurgence during and after the coronavirus disease pandemic, yet access to appropriate care remains limited and disparities persist across regions [1–5]. Globally, one in eight people, approximately 970 million, live with a mental disorder [6, 7], accounting for about 32.4% of years lived with disability (YLDs) and 13.0% of disability-adjusted life years (DALYs) worldwide [8]. The burden is particularly pronounced in sub-Saharan Africa, where prevalence estimates suggest that 3.3–9.8% of the population experiences mood disorders and 5.7–15.8% experiences anxiety disorders. Yet up to 85% of those affected receive no treatment [9]. This lack of appropriate mental health services creates ripple effects across communities, exacerbating untreated conditions, straining fragile health systems, and deepening social and economic disparities [10–16]. Access to professional mental health services remains severely constrained in the region, with the median workforce, including psychiatrists, nurses, psychologists, and social workers, at only 1.6 per 100,000 people, compared with a global median of more than 13 per 100,000 [17]. This paucity of services is further aggravated by stigma surrounding mental illness, which discourages individuals from seeking care and leaves many conditions untreated, increasing pressure on already fragile health systems [16]. Consequently, there is an urgent need to implement cost-effective and accessible mental health delivery systems that can reach underserved populations, reduce barriers to care, and strengthen the resilience of health infrastructures.

Of note, the advent of social media platforms such as Facebook, WhatsApp, X, YouTube, TikTok, and other digital media has significantly transformed communication dynamics[18–23], thus creating new avenues for rapid information dissemination, peer engagement, and interactive dialogue [24–28]. Unlike traditional health communication channels, social network sites and platforms enable users not only to receive information but also to actively engage in discussions about mental health, share personal experiences, and seek guidance from peers and professionals [27]. By harnessing the extensive reach of social media, individuals can access mental health resources and peer support networks in ways that traditional systems often cannot, overcoming geographic, social, and economic barriers to care [29, 30]. While social media usage can accelerate mental health literacy, promote open communication, and encourage help-seeking behaviors by normalizing conversations around psychological well-being, it also offers opportunities for tailored interventions [29, 31–33]. For instance, social media platforms enable targeted awareness campaigns [34], psychoeducational content [35], and peer-led support groups [36] adapted to diverse populations [34–36]. Their interactive nature allows for real-time feedback, helping health communicators to evaluate engagement [37], track behavioral responses [38, 39], and refine messaging strategies to maximize effectiveness and reach [37–39]. However, these benefits are accompanied by potential drawbacks, including the spread of misinformation, exposure to negative or triggering content, and unequal access, which may exacerbate existing disparities in mental health support [40–45].

Furthermore, the potential of social media to shape mental health perceptions is particularly significant, especially in SSA, which has an estimated 384 million users, a figure that continues to grow with expanding internet connectivity and mobile penetration [46]. WhatsApp and Facebook are among the most widely used platforms across the region, with Instagram, YouTube, and TikTok also gaining substantial traction, particularly among youth populations [47]. This uptake unfolds within diverse cultural contexts where traditional healing practices, religious beliefs, and community-based support systems intersect with modern healthcare [48, 49]. These dynamics strongly influence how mental health messages on social media are interpreted, trusted, or resisted, making it essential to align messaging strategies with local cultural realities [50]. Positive narratives on these platforms can improve attitudes toward services and encourage help-seeking [29, 51]; however, poorly framed or misleading content may reinforce harmful stereotypes or spread misinformation [52, 53]. Indeed, some stigma-reduction campaigns, when inappropriately designed, have unintentionally deepened negative perceptions [54–56]. For instance, fear-based HIV campaigns in South Africa, while aiming to promote safer behaviors, reinforced stigma toward people living with HIV rather than reducing it [57]. Similarly, a study in Nigeria shows that framing mental illness in terms of dangerousness or supernatural explanations can worsen public stigma and discourage care-seeking [58]. These insights highlight the importance of considering not only the extent of social media use but also the design, framing, and reception of messages [59, 60]. Consistent with these views, framing theory suggests that the way messages are presented influences how audiences interpret and respond [61], while narrative persuasion theory emphasizes how stories and testimonies facilitate empathy, identification, and behavioral change [62]. Yet when messages are misaligned with cultural expectations, they can yield unintended consequences such as reinforcing stigma or mistrust [48, 63, 64]. In Sub-Saharan Africa, where cultural and community-based frameworks strongly shape mental health perceptions [50, 65], these theories are especially relevant for ensuring that social media campaigns encourage help-seeking rather than harm. Recognizing these dynamics, particularly the interaction between framing, narratives, and cultural expectations, is therefore crucial for developing culturally sensitive digital interventions [49, 66].

Despite the relevance of social media to shape mental health awareness and help-seeking, evidence from SSA remains scarce. Existing studies and/or systematic reviews have largely emphasized prevalence [10, 67–69], barriers to care [70, 71], focused on Western contexts [72], narrow groups, or specific interventions such as mobile apps and SMS campaigns [73–75]. In SSA, few studies have examined how social media messaging itself, through its design, audience characteristics, or unintended effects, influences mental health knowledge, attitudes, behaviors, and help-seeking [51, 72, 76]. This systematic review aims to (1) map the use of social media–based mental health messaging in sub-Saharan Africa and (2) evaluate the impact of these messages on psychological and behavioral outcomes, with particular attention to help-seeking behaviors. The findings will provide evidence to guide the development of culturally relevant digital interventions and strengthen strategies for mental health communication in sub-Saharan Africa.

## Materials and methods

The current review protocol, registered in the International Prospective Register for Systematic Reviews (PROSPERO ID: CRD420251151310), has been prepared following the PRISMA-P checklist (**Supplementary Table 1**) to ensure methodological rigor and transparency [77, 78]. The final review will adhere to PRISMA guidelines [79, 80] and the Cochrane Handbook to ensure robustness and reproducibility [81]. This review aims to synthesize evidence on the effect of mental health messaging on social media platforms (e.g., WhatsApp, Facebook, Instagram, Twitter/X, YouTube, TikTok,) on outcomes such as knowledge (e.g., awareness of symptoms and services), attitudes (e.g., reductions in stigma), empowerment (e.g., confidence in managing stress), behaviors (e.g., adoption of coping strategies), and help-seeking (e.g., intention or action to consult a professional) among populations in SSA. Messaging examples may include Facebook campaigns, WhatsApp-based health groups, or TikTok videos promoting resilience. Online communities and peer-support forums will be included if the mental health messaging is disseminated via social media platforms, but excluded if they exist only as stand-alone websites or private forums.

Additionally, it will compare outcomes between individuals exposed to mental health messaging via social media and those with no exposure or alternative messaging approaches, including variations in framing, content type, or platform (more information about specific interventions and outcomes can be found in Table 1). The protocol outlines the planned methodology, including the search strategy, study selection criteria, data extraction, risk of bias assessment, and synthesis approach. Study selection and database screening will be systematically documented using a PRISMA flow diagram (**Figure 1**). By following a pre-registered protocol, this review ensures that methodological decisions are pre-specified, reducing the risk of reporting bias and enhancing transparency in the evaluation of social media interventions for mental health in SSA.

**Figure 1.**
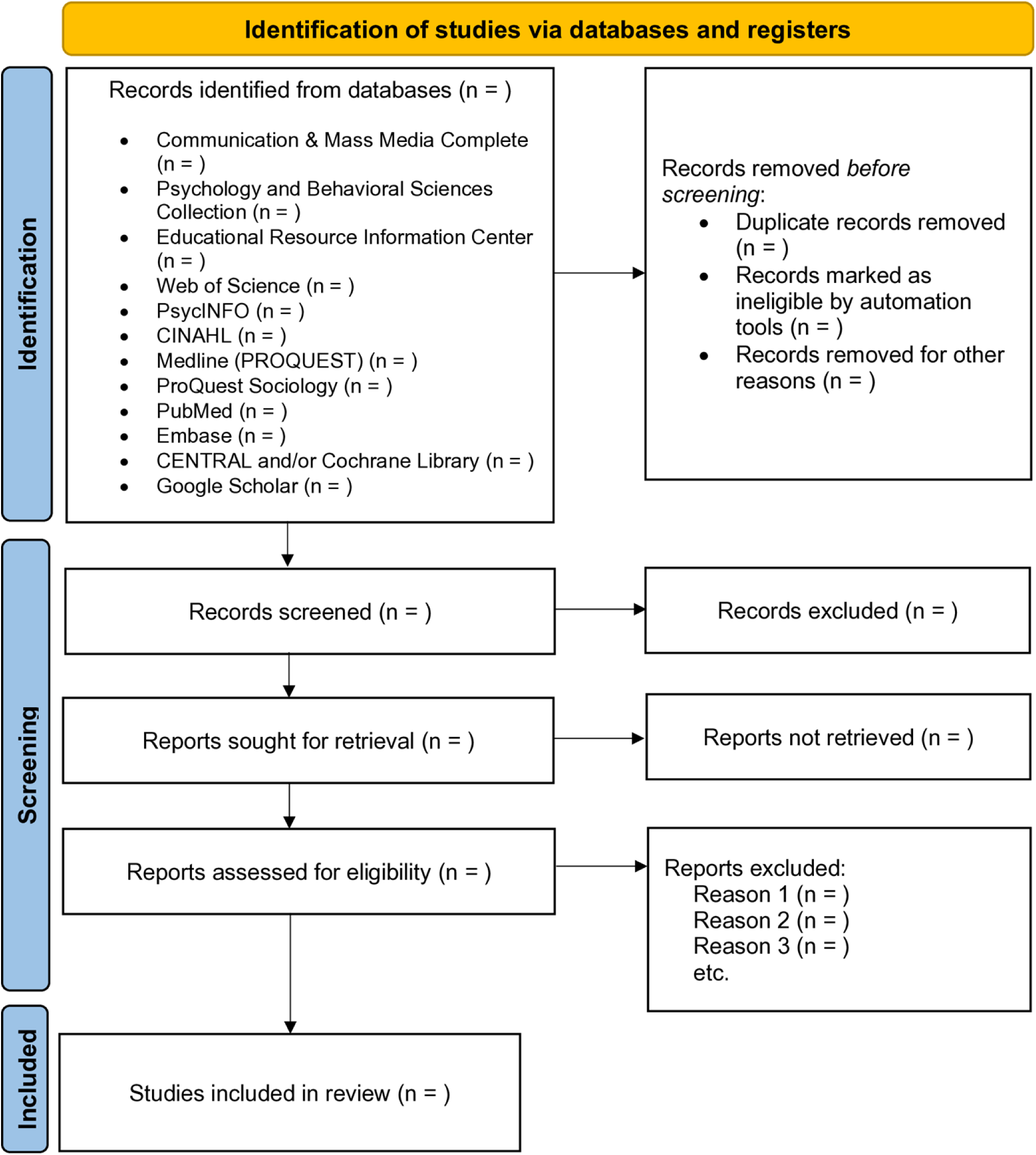
PRISMA flow diagram illustrating the study selection process.

**Table 1:** Search terms adapted for all databases.

| **PICOS Element** | **Description / Search Terms** |
| --- | --- |
| **Population (P)** | Individuals in sub-Saharan Africa (SSA), all countries in SSA, the region as a whole, continental Africa, or general African populations |
| **Intervention / Exposure (I)** | Social media or digital communication: social media, social networks, digital platforms, online platforms, internet, internet-based, web-based, online communication, online community, online network, digital communication, virtual platforms, cyberspace, mobile health (mHealth), eHealth, telehealth, telemedicine, Facebook, WhatsApp, Instagram, YouTube, TikTok, X, Messenger, Telegram, Snapchat, LinkedIn, WeChat, Viber, IMO, Ayoba, ShareChat, Pinterest, Reddit, Threads, , Clubhouse, Google+, MySpace<br>combined with mental health–related terms such as mental health, mental illness, well-being, psychological well-being, psychological distress, psychiatric disorder, mental disorder, behavioral health, major depressive disorder, dysthymia, bipolar disorder, panic disorder, social anxiety disorder, obsessive-compulsive disorder, post-traumatic stress disorder, PTSD, acute stress disorder, adjustment disorder, eating disorder, anorexia nervosa, bulimia nervosa, binge eating disorder, schizophrenia, schizoaffective disorder, psychosis, psychotic disorder, delusion, personality disorder, borderline personality disorder, antisocial personality disorder, narcissistic personality disorder, attention deficit hyperactivity disorder, ADHD, autism spectrum disorder, substance use disorder, alcohol use disorder, drug use |

| PICOS Element | Description / Search Terms |
| --- | --- |
|  | disorder, addiction, gambling disorder, sleep disorder, insomnia, circadian rhythm disorder, burnout, stress, suicidal ideation, suicide, self-harm, self-esteem, mindfulness, emotional regulation, loneliness, social support, quality of life, life satisfaction, happiness, flourish, positive psychology |
| **Comparison (C)** | No exposure, alternative messaging approaches, or standard practice (if applicable) |
| **Outcomes (O)** | **Primary:** Help-seeking (intentions to seek professional support, use of formal mental health services, engagement with informal support networks)<br>**Secondary:** Mental health knowledge (mental health disorders, treatment options, prevention, coping strategies, self-care), attitudes (stigma reduction, acceptance, openness, positive perceptions of help-seeking), behaviors (self-care, treatment adherence, lifestyle changes), empowerment (perceived ability to manage mental health, confidence, self-efficacy), engagement metrics (message reach, interaction rates, self-reported engagement) |
| **Study Design (S)** | All study designs; no restrictions on language or date. Searches will include controlled vocabulary and free-text terms; Boolean operators and truncation will be applied; strategies adapted per database (PubMed / MEDLINE, PsycINFO, CINAHL, CMMC, Scopus, WoS, ERIC, ProQuest Soc., Medline (ProQuest), Embase) |

PubMed/MEDLINE – Medical Literature Analysis and Retrieval System Online (via PubMed), PsycINFO – Psychological Information Database, CINAHL – Cumulative Index to Nursing and Allied Health Literature, CMMC – Communication & Mass Media Complete, Scopus – Scopus, WoS – Web of Science Core Collection, ERIC – Education Resources Information Center, ProQuest Soc. – ProQuest Sociology, Medline (ProQuest) – MEDLINE via ProQuest, Embase – Excerpta Medica Database.

### Eligibility Criteria

#### Inclusion

The criteria for inclusion will follow the PICOS elements, which comprise the population, intervention, comparators, outcomes, and study design.

#### Population (P)

Studies will be eligible if they examine populations residing in sub-Saharan Africa, including adolescents, young adults, adults, and older adults. Countries considered within the review include Angola, Benin, Botswana, Burkina Faso, Burundi, Cabo Verde, Cameroon, Central African Republic, Chad, Comoros, Congo, Côte d’Ivoire, Democratic Republic of the Congo, Djibouti, Equatorial Guinea, Eritrea, Eswatini, Ethiopia, Gabon, Gambia, Ghana, Guinea, Guinea-Bissau, Kenya, Lesotho, Liberia, Madagascar, Malawi, Mali, Mauritania, Mauritius, Mozambique, Namibia, Niger, Nigeria, Rwanda, São Tomé and Príncipe, Senegal, Seychelles, Sierra Leone, Somalia, South Africa, South Sudan, Sudan, Tanzania, Togo, Uganda, Zambia, and Zimbabwe. Studies including multiple regions will be eligible if results specific to sub-Saharan African populations are reported separately.

#### Intervention (I)

The intervention of interest is mental health messaging delivered via social media platforms, including campaigns, posts, videos, advertisements, educational content, digital storytelling, narrative interventions, or other content aimed at improving mental health literacy, reducing stigma, promoting empowerment, or encouraging help-seeking behaviors. Eligible social media platforms include Facebook, WhatsApp, X, Instagram, TikTok, YouTube, Snapchat, Pinterest, LinkedIn, Telegram, Messenger, WeChat, Viber, IMO, Ayoba, ShareChat, Reddit, Threads, Clubhouse, Google+, and MySpace. Messaging may vary in format, frequency, duration, and framing, including persuasive communication, narrative storytelling, risk communication, or public health education.

#### Comparator (C)

When available, comparators include populations with no exposure to mental health messaging or exposure to alternative messaging approaches, such as different message framing, delivery platforms, content types, or campaign intensity.

#### Outcome (O)

The primary outcome of interest is help-seeking, encompassing intentions to seek professional support, utilization of formal mental health services, and engagement with informal support networks. Secondary outcomes include mental health–related knowledge, attitudes, behaviors, empowerment, and engagement. Specifically, knowledge outcomes cover understanding of mental health disorders, treatment options, prevention, coping strategies, and self-care practices. Attitudes involve stigma reduction, acceptance of mental health issues, openness to discussion, and positive perceptions of help-seeking. Behavioral outcomes include engagement in self-care, adherence to treatment, and lifestyle changes that promote well-being. Empowerment outcomes reflect perceived ability to manage mental health, confidence in seeking support, and self-efficacy. Other outcomes include engagement metrics such as message reach, interaction rates, or self-reported interaction with content.

#### Study design (S)

Eligible study designs include randomized controlled trials, cluster-randomized trials, quasi-experimental studies such as pre-post designs or interrupted time series, cohort studies, cross-sectional surveys, qualitative studies (e.g., interviews, focus groups, ethnographies) and mixed-methods studies examining exposure to social media mental health messaging.. Both quantitative and qualitative outcomes related to mental health messaging will be included.

### Exclusion

Studies not reporting relevant mental health outcomes, not conducted in sub-Saharan Africa, or focusing solely on offline interventions will be excluded. Reviews, editorials, commentaries, conference abstracts without full-text data, and studies not published in English or French will be excluded.

### Data sources

Electronic databases to be systematically searched include PubMed/MEDLINE ((Medical Literature Analysis and Retrieval System Online) via PubMed), PsycINFO (Psychological Information Database), CINAHL (Cumulative Index to Nursing and Allied Health Literature), Communication & Mass Media Complete, Scopus, Web of Science Core Collection, ERIC, ProQuest Sociology, Medline via ProQuest, and Embase if accessible. Searches will cover studies from January 2000 to September 2025. In addition, hand searches will be conducted on the first ten pages of Google Scholar, and reference lists of included studies and relevant reviews will be reviewed manually. Key journals focusing on global mental health, digital health interventions, and social media studies will also be reviewed for potentially relevant studies not indexed in the electronic databases.

### Search terms, and search strategies for identifying studies

Searches will be conducted in the aforementioned databases using four main concepts aligned with the PICOS framework (**Table 1**): mental health terms, messaging or communication terms, social media or digital terms, and population terms. The search will cover studies published between January 2000 and September 2025, a period that reflects the rise and widespread use of social media. Only studies in English will be included. Boolean operators and truncation will be applied to combine synonyms and variations, and the strategy will be adapted for each database using controlled vocabulary and free-text terms. Mental health terms include, but are not limited to, mental health, mental illness, wellbeing, psychological well-being, psychological distress, psychiatric disorders, affect, emotion, depression, major depressive disorder, dysthymia, bipolar disorder, anxiety, panic disorder, phobias, social anxiety disorder, obsessive-compulsive disorder, post-traumatic stress disorder, acute stress disorder, adjustment disorder, eating disorders, schizophrenia, psychosis, personality disorders, attention deficit hyperactivity disorder, autism spectrum disorders, substance use disorders, alcohol and drug use disorders, addiction, gambling disorder, sleep disorders, burnout, stress, suicidal ideation, self-harm, resilience, coping, self-esteem, mindfulness, emotional regulation, loneliness, social support, quality of life, life satisfaction, happiness, flourishing, and positive psychology. Messaging or communication terms include message, communication, health promotion, health campaign, public health message, media message, mental health communication, intervention, health education, information, content, outreach, digital health communication, risk communication, social marketing, online message, persuasive communication, narrative, storytelling, framing, health information, information dissemination, knowledge translation, and education. Social media and digital terms include social media, social networks, digital platforms, online platforms, internet, internet-based, web-based, online communication, online community, online network, digital communication, virtual platforms, cyberspace, mobile health, mHealth, eHealth, telehealth, telemedicine, Facebook, WhatsApp, Instagram, YouTube, TikTok, X, Messenger, Telegram, Snapchat, LinkedIn, WeChat, Viber, IMO, Ayoba, ShareChat, Pinterest, Reddit, Threads, Clubhouse, Google+, and MySpace. Population terms include all countries in sub-Saharan Africa, the region as a whole, continental Africa, or general African populations. Full search strategies will be documented in the review report for transparency and reproducibility.

### Study Selection

All records retrieved will be exported to EndNote reference management software to automatically exclude any duplicates. Subsequently, the references will be exported to the Covidence platform for titles, abstract screening, and full text review. Specifically, two independent reviewers (P.A.B. and J.A.) will screen titles and abstracts for eligibility based on the a priori eligibility criteria. A Cohen’s kappa will be calculated as a measure of intercoder reliability. Discrepancies will be resolved through discussion, and a third reviewer will adjudicate unresolved disagreements. Full-text articles will then be screened independently by two reviewers according to the pre-specified inclusion and exclusion criteria, and reasons for exclusion at the full-text stage will be documented in a PRISMA flow diagram.

### Data Extraction

Data will be extracted using a standardized form developed and piloted prior to full extraction. Extracted information will include study characteristics such as author, year, country, study design, and setting; population characteristics including sample size, age, gender, urban or rural context, socioeconomic status, and other relevant information (e.g., community sample, clinical sample, etc.); intervention characteristics including social media platform, message type, duration, frequency, framing, and content type; comparator characteristics; outcomes including type, conceptualization, measurement tool, timepoints, and reported effects; key findings, including quantitative results (and, when possible, effect sizes and confidence intervals) and qualitative findings. Two reviewers (P.A.B. and J.A.) will extract data independently, and discrepancies will be resolved through consensus.

### Risk of Bias

The methodological quality and risk of bias of included studies will be assessed independently by two reviewers. Randomized controlled trials will be evaluated using the Cochrane Risk of Bias 2 (RoB 2) tool. At the same time, all observational and non-randomized studies (including cross-sectional, cohort, case-control, and quasi-experimental designs) will be appraised using the STROBE checklists to ensure consistency and comparability across study types.

### Quality Assessment

To provide a comprehensive evaluation of study quality, the National Heart, Lung, and Blood Institute (NHLBI) Study Quality Assessment Tools will be employed. These tools are specifically designed to assess internal validity, potential sources of bias, and overall methodological rigor across different study designs, including controlled intervention studies, cohort and cross-sectional studies, case-control studies, and before-after studies. Each study will be rated as good, fair, or poor quality based on criteria such as clarity of research question, adequacy of study population and sample size, appropriateness of exposure or intervention measurement, outcome assessment, statistical analyses, and control for confounding variables. The combination of risk of bias assessment tools and the NHLBI quality appraisal ensures a thorough evaluation of the credibility and reliability of the evidence. Discrepancies in quality assessment will be resolved through discussion or consultation with a third reviewer (I.O.D.J. or L.M), and the quality ratings will be incorporated into the narrative synthesis and interpretation of findings.

### Data Synthesis

For heterogeneous data, we plan to narratively consolidate the evidence by synthesis without meta-analysis [82]. Specifically, the narrative synthesis will be conducted due to anticipated heterogeneity in study designs, intervention types, platforms, and outcomes. The synthesis will be structured around population characteristics, intervention characteristics including platform, messaging type, duration, and framing, comparator type, and mental health outcomes including knowledge, attitudes, behaviors, empowerment, and help-seeking [82]. Where feasible, meta-analysis will be conducted using random-effects models if at least two studies are sufficiently homogeneous in terms of population, intervention, and outcomes. The *metaprop function* in R (from the *meta* package) will be used for pooling proportions and estimating overall effects. [83].

Effect sizes, including standardized mean differences or odds ratios, will be calculated with 95% confidence intervals. Statistical heterogeneity will be assessed using the I² statistic, with values of 25 percent (%), 50%, and 75% interpreted as low, moderate, and high heterogeneity, respectively. Subgroup analyses may be conducted based on age group, social media platform, message framing, or urban versus rural setting. Sensitivity analyses will be conducted by excluding studies with a high risk of bias.

### Publication Bias

Publication bias will be assessed through visual inspection of funnel plots if at least ten studies are included in a meta-analysis. Egger’s test or Begg’s test may also be conducted to quantitatively assess the presence of small-study effects.

### Confidence in Cumulative Evidence

The Grading of Recommendations Assessment, Development, and Evaluation (GRADE) approach will be used to assess the certainty of evidence across studies for each outcome. This involves evaluating the body of evidence based on five domains: risk of bias, inconsistency of results, indirectness of evidence, imprecision, and publication bias. Based on these domains, evidence will be rated as high, moderate, low, or very low based on considerations of risk of bias, consistency, directness, precision, and publication bias. Applying GRADE will ensure that the review not only summarizes findings but also conveys the strength and reliability of the evidence, which is critical for informing culturally relevant interventions in sub-Saharan Africa.

### Handling of Missing Data

In instances where included studies have missing, incomplete, or unclear data, efforts will be made to contact the corresponding authors to request additional information or clarification. When missing data cannot be obtained, the extent and nature of missing information will be documented and reported. For quantitative outcomes, the potential impact of missing data on study findings will be considered, and sensitivity analyses will be conducted when feasible to assess how assumptions about missing data affect the results. Studies with substantial or critical missing data that could compromise the validity of outcomes may be rated as lower quality or excluded from meta-analysis, while their findings will still be included in the narrative synthesis. The presence of missing data will be taken into account during the risk of bias assessment and quality appraisal using the Cochrane Risk of Bias 2 tool, ROBINS-I, and STROBE checklist. By systematically documenting and addressing missing data, the review aims to minimize potential bias and ensure that conclusions are based on the most reliable and complete evidence available.

### Ethics and Dissemination

As this review will synthesize evidence from previously published studies, ethical approval is not required. Findings will be disseminated through peer-reviewed publication, presentations at international conferences, and policy briefs aimed at stakeholders involved in mental health promotion in sub-Saharan Africa.

## Discussion

The proposed systematic review addresses a critical knowledge gap in understanding the role of social media in shaping mental health awareness, attitudes, and help-seeking behaviors in SSA. While social media platforms have been shown to facilitate rapid information dissemination, peer engagement, and interactive support globally, evidence from SSA remains limited. Most existing studies in the region have primarily focused on mental health prevalence [10, 67–69], barriers to care [70, 71], or the evaluation of specific digital interventions such as mobile applications and SMS campaigns [73, 84] in Western countries. Few studies have examined how the design, framing, and audience characteristics of social media messaging directly influence mental health literacy, stigma reduction, and help-seeking behaviors in SSA [72, 85–87]. By synthesizing available evidence, this review aims to clarify how social media messaging can be leveraged to promote culturally sensitive and effective mental health interventions, particularly in a context where traditional support systems and community norms intersect with modern healthcare delivery.

Despite its potential contributions, several limitations are anticipated in this review. First, the heterogeneity of study designs, social media platforms, messaging formats, and outcome measures may limit the ability to draw generalized conclusions. Second, the rapidly evolving nature of social media use and digital health communication in SSA means that findings may quickly become outdated. Third, potential publication bias and the limited availability of peer-reviewed studies from low-resource settings could restrict the comprehensiveness of the evidence base. Finally, the review may be constrained by the quality and reporting of included studies, particularly regarding cultural adaptation, message framing, and the measurement of behavioral outcomes. Nonetheless, by systematically identifying and evaluating the current literature, this review will provide critical insights into digital mental health communication strategies and inform the design of interventions that are both evidence-based and culturally relevant for SSA populations.

## Supporting information

S1 File. PRISMA-P (Preferred Reporting Items for Systematic Review and Meta-Analysis Protocols) 2015 checklist: Recommended items to address in a systematic review protocol.

## Abbreviation

CENTRAL — Cochrane Central Register of Controlled Trials, CINAHL — Cumulative Index to Nursing and Allied Health Literature, DALYs — disability-adjusted life years, eHealth — electronic health, ERIC — Education Resources Information Center, GRADE — Grading of Recommendations Assessment, Development, and Evaluation, I² — I-squared statistic (measures between-study heterogeneity in meta-analysis), ID — Identifier, JBI — Joanna Briggs Institute, MEDLINE — Medical Literature Analysis and Retrieval System Online, mHealth — mobile health, NHLBI — National Heart, Lung, and Blood Institute, PICO — Population, Intervention, Comparator, Outcome, PICOS — Population, Intervention, Comparator, Outcomes, Study design, PRISMA — Preferred Reporting Items for Systematic Reviews and Meta-Analyses, PRISMA-P — Preferred Reporting Items for Systematic Review and Meta-Analysis Protocols, PROSPERO — International Prospective Register of Systematic Reviews, PsycINFO — American Psychological Association (APA) database, RoB 2 — Cochrane Risk of Bias 2, ROBINS-I — Risk of Bias in Non-randomized Studies of Interventions, SMS — short message service, SSA — sub-Saharan Africa, STROBE — Strengthening the Reporting of Observational Studies in Epidemiology, YLDs — years lived with disability

## Authors contribution

**Conceptualization:** P.A.B., I.O.D.J. and L.M.; **Project administration:** P.A.B., I.O.D.J., J.A., M.D.H, and L.M; **Resources**: P.A.B., I.O.D.J., J.A., M.D.H, and L.M; **Methodology:** P.A.B., I.O.D.J., J.A., M.D.H, and L.M; **Investigation:** P.A.B., I.O.D.J., J.A., M.D.H, and L.M; **Software:** P.A.B., I.O.D.J., J.A., M.D.H, and L.M; **Writing original draft:** P.A.B., I.O.D.J., J.A., M.D.H, and L.M; **Writing reviews and editing:** P.A.B., I.O.D.J., J.A., M.D.H, and L.M; **Correspondence:** L.M; **Supervision:** L.M.

## Data Availability

No new data were generated for this study. All information relevant to the protocol is included within the manuscript and its supplementary information files.

## Ethical Consideration

This study is a protocol using secondary sources; hence, ethical approval was not required.

## Acknowledgements

None

## Consent for Publication

Not applicable.

